# Quantitative analysis of the iris surface smoothness by spectral domain optical coherence tomography in Fuchs uveitis

**DOI:** 10.1101/2020.05.06.20093302

**Authors:** Mohammad Zarei, Elias Khalili Pour, Nazanin Ebrahimiadib, Hamid Riazi-Esfahani

**Affiliations:** Retina Service, Farabi Eye Hospital, Tehran University of Medical Sciences, Tehran, Iran, Qazvin Square, South Kargar Street, Tehran 1336616351, Iran

**Keywords:** Fuchs uveitis, Iris atrophy, Smoothness index, Anterior segment OCT

## Abstract

**Purpose:** To demonstrate the value of “smoothness index” as a novel tool for quantitative analysis of iris surface findings in unilateral Fuchs’ uveitis (FU).

**Methods:** In this observational study, both phakic eyes from patients with the diagnosis of unilateral FU were enrolled. All eyes underwent a complete ophthalmic examination and anterior segment optical coherence tomography(AS-OCT). The images were extracted and exported to the ImageJ software for calculating the “smoothness index (SI)”. The SI was defined as ratio of length of the straight line connecting the most peripheral and the most central points of anterior iris surface to actual length of this boundary.

**Results:** Forty eyes from 20 patients with unilateral FU were enrolled. Mean age of patients was 40±10 years. Mean overall SI was 0.876± 0.030 in eyes with FU that was significantly more than fellow control eyes (0.841± 0.041). (p=0.005) The mean temporal and nasal SI were 0.878± 0.035 and 0.874± 0.033 in eyes with FU, compared to 0.843± 0.041 and 0.838± 0.047 in control fellow eyes, respectively. (p = 0.008 and p = 0.009, respectively)

**Conclusion:** Iris “smoothness index” is a novel index to quantitatively document diffuse smoothness of iris anterior surface in eyes with FU. This index may facilitate diagnosis of subtle iris atrophic changes in these eyes, especially when heterochromia is absent.

## Introduction

Fuchs Uveitis (FU) is a chronic, often unilateral ocular inflammation that mainly affects anterior uvea.[1] The diagnosis is primarily based on clinical findings which can vary among different ethnicities. [2] It is estimated that 1–6% of patients who are referred to uveitis clinics are diagnosed with FU.[3] Presenting symptoms are usually subtle. Most cases manifest with characteristic fine stellate keratic precipitates (Fuchs KPs), mild anterior chamber reaction, iris atrophic changes, vitreous cells and debris, posterior subcapsular cataract, glaucoma, absence of macular edema, and absence of posterior synechiae.[4, 5] Despite the well-known clinical picture, incorrect diagnosis of FU is still a frequent problem.[4, 5]

No imaging modality can definitely confirm the diagnosis of FU. However, some features in fluorescein angiography (FA) such as disc hyperfluorescence and absence of angiographic cystoid macular edema (CME) are suggestive.[6, 7]

Heterochromia, has been historically considered as one of the most characteristic findings of FU. It has been proposed that thinning and pigment loss in all iris layers, including the iris pigment epithelium, stroma and anterior border lead to heterochromia.[2, 8, 9] Reported frequency of heterochromia varies widely between populations. While it is a prominent feature in patients from Western European descent, it is not a frequent finding in patients with dark eyes.[4, 10, 11] Therefore, in the latter group, focusing on heterochromia as a key to diagnose FU can be misleading. However, diffuse atrophic changes of iris other than heterochromia, seems to be much more common in patients from these populations and searching for these subtle changes may lead to correct diagnosis. With the advent of anterior segment optical coherence tomography (AS-OCT), some investigators have tried to quantitatively document iris atrophy in FU eyes with conflicting results.[5, 12, 13] Some concluded that iris stromal thickness may not be a sensitive representative of iris atrophic changes in this syndrome.[13]

Among the clinical iris atrophic findings of FU, the presence of a relatively featureless or cryptless iris is especially helpful: on slit lamp examination, anterior surface of the iris seems to have less crypts than a normal eye. This diffuse “smoothness” of anterior iris surface of the affected eye is usually in sharp contrast with anterior iris surface of the healthy fellow eye in unilateral cases.[5] In our practice, we found this sign to facilitate the correct diagnosis in challenging cases. We have previously shown that in AS-OCT images of patients with unilateral FU, the smoother iris of affected eye can be recognized from the healthy fellow eye by masked observers. One potential drawback of this method is its qualitative nature. To address this deficit, we have proposed a method to calculate the “smoothness index” in AS-OCT images.[5] The objective of the current study is to evaluate the usefulness of this index in a group of patients with unilateral FU.

## Materials and methods

This observational study was conducted at Farabi Eye Hospital, Tehran, Iran. The study followed the tenets of the Declaration of Helsinki and was approved by the local Institutional Review Board (IRB).

Both phakic eyes from patients with the diagnosis of unilateral FU entered the study. Diagnosis of unilateral FU was made by either of two authors (M.Z. and N.E) and was based on the presence of chronic anterior uveitis, presence of typical Fuchs KPs, absence of posterior synechiae, absence of macular edema in slit lamp examination, and absence of macular edema in macular spectral domain OCT (SD-OCT) and/or FA. Presence of posterior subcapsular cataract, open angle glaucoma or ocular hypertension, vitreous cells or degenerative changes, dull appearance of iris surface (compared to healthy fellow eye), and heterochromia were considered supportive, but not necessary for the diagnosis of FU.

Patients who were using any medication affecting pupillary diameter, cases with intumescent cataract, and patients with a history of ocular trauma, laser treatment or surgeries in either eye were excluded. Patients with any abnormal focal or multifocal findings in iris structure (other than nodules) were also excluded.

All patients underwent a complete ophthalmic exam in both eyes in addition to AS-OCT scanning (swept source (SS)-OCT CASIA 1 or 2, Tomey, Japan) of both eyes in the same session, in regular day room illumination without prior administration of any mydriatic or miotic drops.

Patients underwent horizontal AS-OCT B scans coursing the iris from 3 o’clock to 9 o’clock passing the pupillary center. During the imaging, patients were instructed to open their eyes as wide as possible and not to blink. If any artefact was noted, the imaging was repeated until an acceptable image was achieved. Images were exported to and analysed with ImageJ (ImageJ version 1.52, NIH, USA) software. All images were manually segmented and measured by one author (E.K) who was masked to the affected eye. To measure the lengths, the free-hand tool of ImageJ with 300 percent magnification was used.

As we previously proposed [5], smoothness index (SI) was defined as the ratio of the length of the straight line connecting the most peripheral and the most central points of anterior iris surface (in nasal and temporal sides) to actual length of this boundary (in nasal and temporal sides) (Figure 1). To calculate the overall SI, the sum length of nasal and temporal “straight lines” was divided by the sum of nasal and temporal actual lengths of anterior iris boundary.

**Fig. 1.**
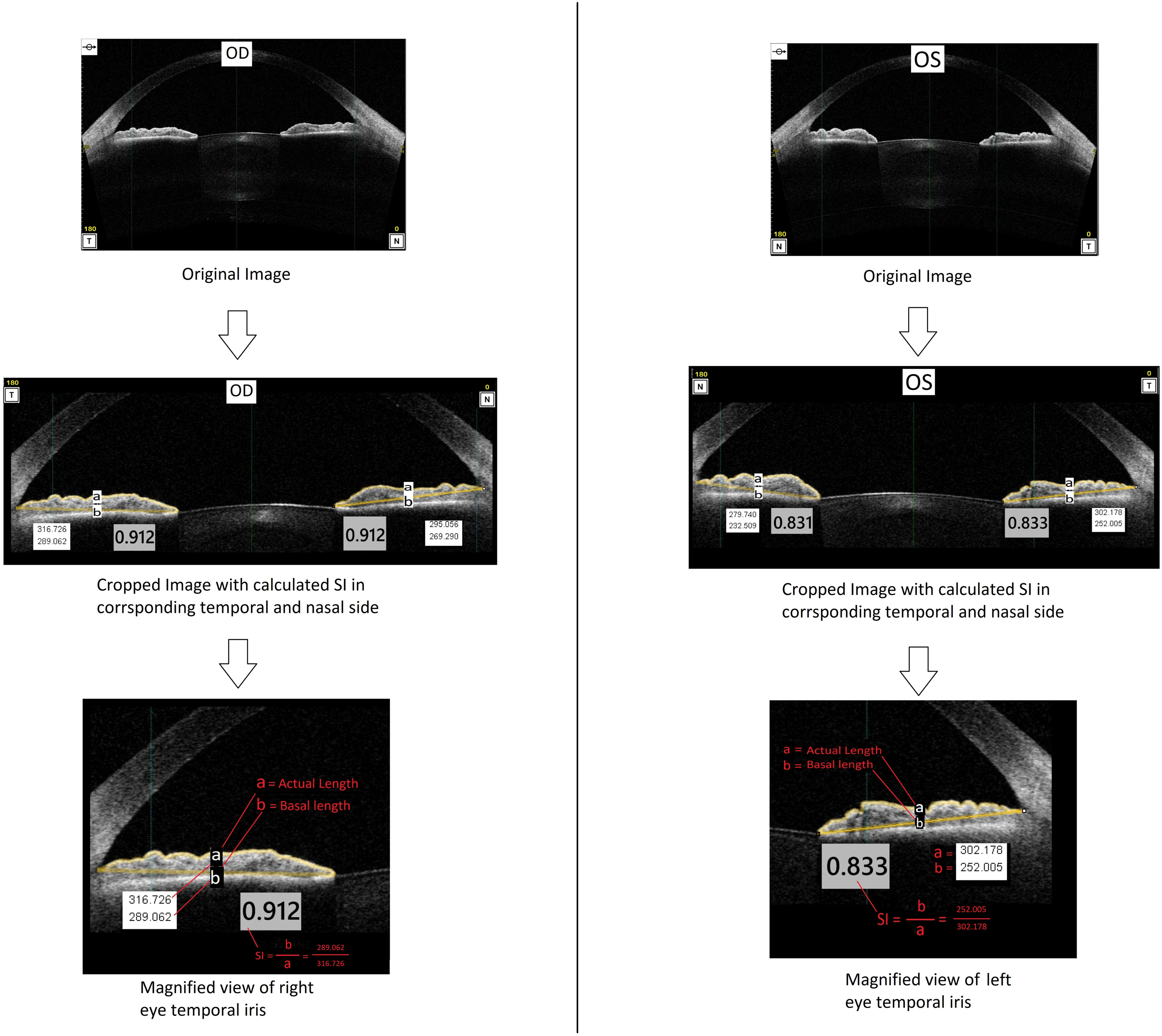
Smoothness index (SI), is the ratio of the length of the straight line connecting the most peripheral and the most central points of anterior iris surface (“b” in nasal and temporal sides) to the actual length of this boundary (“a” in nasal and temporal sides). To calculate the overall SI, the sum length of nasal and temporal “straight lines” is calculated and divided by the sum of nasal and temporal actual lengths of anterior iris boundary. In this case, the right eye is affected by unilateral Fuchs uveitis.

## Statistical analysis

For statistical analysis, SPSS software (IBM Corp. Released 2013. IBM SPSS Statistics for Windows, Version 22.0. Armonk, NY: IBM Corp.) was used and *p* values less than 0.05 were considered significant. To compare corresponding SIs between FU eyes and healthy fellow eyes, linear mixed model (LMM) was used.

## Results

Forty eyes from 20 patients with unilateral FU were enrolled. Mean age of patients was 40±10 (27-69) years. Eleven (55%) patients were males. In eight patients, right eye was affected by FU.

Mean VA (logMAR) in FU eyes and healthy fellow eyes were 0.70± 0.53 (0.00-1.70) and 0.02± 0.07 (0.00-0.30), respectively.

Unilateral chronic anterior uveitis with typical Fuchs KPs was found in all cases. Vitreous cells and vitreous degeneration were found in 15 (75%) and 10 (50%) of FU eyes, respectively. Posterior subcapsular cataract and heterochromia were found in 16 (80%) and two (10%) FU eyes, respectively. Intraocular pressures >21mmHg were found in two (10%) FU eyes and none of fellow eyes.

Mean temporal SI was 0.878± 0.035 (range= 0.793-0.917) and 0.843± 0.041 (range= 0.770-0.928) in FU eyes and fellow eyes, respectively (estimated mean difference = −0.034, 95% CI [−0.058, −0.009], p=0.008). Mean nasal SI was 0.874± 0.033 (range= 0.803-0.920) and 0.838± 0.047 (range=0.755-0.903) in FU eyes and fellow eyes, respectively (estimated mean difference = −0.035, 95% CI [−0.061, −0.009], p=0.009). Mean overall SI was 0.876± 0.030 (range= 0.798-0.91) and 0.841± 0.041 (range= 0.768-0.906) in FU eyes and fellow eyes, respectively (estimated mean difference = −0.034, 95% CI [−0.058, −0.011], p=0.005). Summary of the results is shown in table 1.

## Discussion

Clinical iris changes in FU have been studied extensively.[8, 14-17] These changes include iris nodules, refractile Russell bodies, heterochromia, abnormalities in iris vessels, a smooth iris surface with a “washed-out” appearance, absent or interrupted pupillary pigment ruff, and transillumination defects in posterior pigment epithelium. These signs are best appreciable in comparison to healthy fellow eye in unilateral cases. Posterior synechiae is characteristically absent. Most of these iris alterations have been attributed to iris atrophy and are characteristically diffuse; so that localized or sectoral changes are strongly in favour of diagnoses other than FU (e.g. herpetic uveitis). The atrophy can involve all iris layers (anterior border layer, stroma, and posterior pigment epithelium), however a predilection towards more anterior layers has been suggested.[17] It is also proposed that smooth iris surface with dull stroma and transillumination defects in posterior pigment epithelium are more prominent in areas adjacent to pupil. [16, 17]

It has been suggested that stromal iris atrophy may highlight some deeper structures of iris (vasculature and sphincter pupillae), especially in advanced cases. However, in most cases this atrophy leads to blunting of surface features and the affected iris appears to be smoother than the healthy fellow eye.[8, 16, 17]

It is well known that in FU, heterochromia is more prevalent in patients from Western European descent than patients from other ethnic groups.[9, 11, 13, 18] Heterochromia is not a common feature in FU patients in Asia and middle east.[4, 13, 18] However, atrophic iris changes other than heterochromia seems to be common, even in populations not from Western European descent. [11, 18, 19]

With the advent of AS-OCT, investigators tried to quantitatively document these iris atrophic changes in vivo with this novel imaging modality. The usual approach has been the measurement of iris stromal thickness to differentiate the affected eye from normal fellow eye. However, various studies reported contradicting results. Basarir et al. [12] studied iris and anterior chamber structure in eyes affected by FU with time domain anterior segment OCT (Visante OCT 3.0 Model 1000, Carl Zeiss Meditec, Inc). They measured iris thickness in 38 patients with unilateral FU at three locations: at the thickest part of iris, at 500 μm from the scleral spur, and in the middle of the iris. They found that in FU eyes, iris at the thickest part was significantly thinner than the healthy fellow eyes. However, iris thickness at other two locations was not statistically different between FU eyes and healthy fellow eyes. The authors attributed this discrepancy to the presence of crypt troughs in the latter two locations. Diffuse nature of iris atrophy in FU can be another explanation for this finding for which point thickness measurements -as done by Basarir et al- are not optimal representatives.

Later, Invernizzi et al.[20] used the spectral domain OCT (Spectralis OCT Anterior Segment Module (Heidelberg Engineering, Germany)) to measure iris thickness in 14 patients with unilateral FU. They obtained cross-sectional full thickness iris images in four meridians: inferior, temporal, superior and nasal. Instead of measuring iris point thickness, Invernizzi et al aimed to measure mean iris thickness in each meridian. Due to presence of crypts that results in irregular shape of anterior border of iris, Invernizzi et al. introduced a novel method to calculate mean iris thickness: after drawing a 3000 μm horizontal line along the pigmented posterior epithelium from pupillary margin towards iris periphery and importing the image into ImageJ software, they measured the area of the iris section along the aforementioned line. By dividing this value by the 3000 μm, an approximated mean iris thickness through the selected section was obtained. Reported results showed that the iris in the affected eyes was significantly thinner than the healthy fellow eyes. Despite the clear advantage of this method over Basarir et al’s method in obtaining mean iris thickness instead of point thicknesses, there are some considerations with this method. First, because of limited light penetration through highly pigmented layers, posterior iris border appears blurry and difficult to identify in scanned images. To resolve this challenge, the authors decided to exclude posterior pigmented epithelial layer from the section. However, they believed that exclusion of posterior pigment epithelium -consisting of a double layer of cells- did not significantly confound the measurement. Second, using a 3000 μm line, starting from pupillary margin towards the iris periphery, exclude the most peripheral part of iris from the analysis. Third, it is expected that in some cases, iris bowing (anterior convexity) makes it difficult to accommodate a “horizontal” 3000 μm line to the posterior border of the iris which may result in imprecise estimation of the area of selected iris section and erroneous calculation of mean iris thickness. Fourth, it should be noted that from a histopathologic point of view, decreased volume is the most relevant indicator of atrophy. However, measuring the iris volume was not feasible for Invernizzi et al. In this case, compared to iris thickness, the area of iris cross-section provides a more direct assessment of iris atrophy. Therefore, to demonstrate iris atrophy, calculating an approximated mean iris thickness from the measured area of iris cross-section, seems to be unnecessary; a direct comparison of the selected areas between fellow eyes – which were already measured- is preferred.

Ozer et al.[13] used similar machinery and analysis method as Invernizzi et al to measure mean iris thickness in nasal and temporal quadrants of 21 patients with unilateral FU. However, they could not find a statistically significant difference between FU eyes and healthy fellow eyes. They attributed this outcome to the small number of enrolled patients. The authors also pointed to possible effect of racial differences as a conflicting factor in comparison of the results of different studies.

It is noteworthy that prior to the advent of OCT, using the term “atrophy” for description of iris findings in FU, was largely based on observations with slit lamp exam, not on the direct measurement of iris thickness in histopathological studies. To the best of authors’ knowledge, there is no documented histopathologic evidence of decreased iris thickness in English-language ophthalmologic publications.[14, 15, 21] In other words, clinicians inferred that the best explanation for observed iris changes in clinical exam, should be the decreased thickness/volume of iris tissue. However, most of what was observed with slit lamp exam, was actually a decrease in surface features and pigmentation of iris in FU eye compared to healthy fellow eye.[8, 14-17] Therefore, for quantitative documentation of iris changes observed in clinical exam, finding a way to measure smoothness of iris surface maybe more relevant than measuring the iris thickness. As we previously reported, in unilateral FU, careful qualitative examination of AS-OCT images can be very helpful in detection of decreased prominence of iris crypts in FU eyes compared to healthy fellow eyes. This may potentially help the clinician to correctly diagnose dubious cases of FU. However, to be more precise and achieve objective measurements, we defined an AS-OCT-derived quantitative index for assessing smoothness of iris surface. [5] To the best of our knowledge, this study is the first one to use this index for evaluating iris atrophic changes in unilateral FU. As we showed, the SI was significantly greater in eyes with FU than in normal fellow eyes, either in temporal or in nasal part of the iris. This index may be especially useful in diagnosing the cases of unilateral FU without prominent heterochromia. Rather than age-gender-matched subjects, we used the healthy fellow eyes of the same patients as control group to minimize the effect of inter-individual differences in the analysis.

In this study our goal was to evaluate a quantitative OCT counterpart (smoothness index) for iris atrophic findings which are observed in routine eye exam in FU patients. Therefore, in contrast to Invernizzi et al and Ozer et al, we obtained AS-OCT images without pharmacologic miosis and in conditions very similar to conditions under which patients were examined by slit lamp.

A limitation of our study is that all the patients were from the same ethnicity. Usefulness of the smoothness index should be assessed in patients from other populations as well.

Another important limitation is that procedures of export/import and manual segmentation of anterior iris border is time consuming and needs a skillful operator. Development of an automatic or semi-automatic version of this method can make it more feasible in daily practice.

In conclusion, smoothness index is a useful novel index to quantitatively document diffuse smoothness of iris anterior surface in eyes with FU. This index may facilitate diagnosis of subtle iris atrophic changes in these eyes, especially when heterochromia is absent; a frequent scenario in patients from non-Western European descent.

## Data Availability

The data generated during or/and analysed during the current study are available from the corresponding author

